# Detection and quantification of key dental pathogens through wastewater monitoring

**DOI:** 10.1101/2025.07.03.25330793

**Authors:** Olivia N. Birch, Sang C. Par, Justin C. Greaves

## Abstract

Wastewater-based epidemiology (WBE) has been widely used to track viral pathogens like SARS-CoV-2 and polio, but its potential for monitoring common dental bacterial pathogens that infect the oral cavity has yet to be explored. *Streptococcus mutans* and *Porphyromonas gingivalis* are key oral bacterial pathogens that cause highly prevalent dental diseases worldwide, such as dental caries and gingivitis. Our main objective for this study was to investigate the presence and prevalence of these oral bacteria in wastewater to determine the feasibility of using WBE for oral pathogens. We measured *S. mutans* and *P. gingivalis* nucleic acids in weekly samples for 24 months at a local wastewater treatment plant. A total of 100 untreated wastewater samples were collected once a week between June 2023 and May 2025. Samples were concentrated, extracted for DNA, then tested for each bacterium. Our results showed that 89% and 58% were positive for *S. mutans* and *P. gingivalis*, respectively, which shows that wastewater surveillance is appropriate for oral bacteria. Average concentrations were 4.57 log_10_ genome copies/L and 3.03 log_10_ genome copies/L for *S. mutans* and *P. gingivalis*, respectively. Detections of oral bacteria were observed in the primary and final effluent but concentrations were significantly lower in the final effluent than untreated wastewater. The high levels of oral bacteria in wastewater indicated a potential transmission mechanism for these bacteria through water, specifically for *S. mutans*. Additionally, this study underscores the unique potential for WBE to be used in the surveillance of oral bacterial pathogens.

## 1.0 Introduction

Dental caries, or tooth decay, is one of the most prevalent chronic diseases worldwide, affecting nearly 2.3 billion people with permanent tooth decay and over 530 million children with primary tooth loss, according to the World Health Organization (Organization, 2017). *S. mutans* is a major culprit in dental caries, playing a significant role in plaque formation and acid production that demineralizes tooth enamel (Caufield, 2000; Forssten et al., 2010; Lemos et al., 2019). *P. gingivalis*, on the other hand, is a key pathogen in periodontal disease, a severe gum infection that can lead to tooth loss and systemic health complications which can include cardiovascular disease and diabetes (Marutescu et al., 2023; Rafiei et al., 2017). The high global burden of these oral health issues underscores the importance of developing innovative disease surveillance methods to understand the spread of these diseases.

Wastewater Based Epidemiology (WBE) has become increasingly popular in recent years as a tool to monitor the spread of endemic and emerging pathogens passively in a community. WBE has already been successfully employed to monitor various pathogens, such as SARS-CoV-2 and antimicrobial resistance genes, yet its application to oral bacteria remains largely unexplored (Greaves et al., 2020b; Rodriguez et al., 2024). Given the widespread presence of these oral bacteria in human populations, understanding their prevalence in wastewater can provide insights into community-level oral health trends and transmission dynamics. Traditional surveillance methods, such as clinical examinations and self-reported surveys, have limitations in reach, accuracy, and cost-effectiveness (Pitiphat et al., 2002; Sekundo et al., 2019). WBE, on the other hand, allows for large-scale, community-wide monitoring without the need for individual participation, offering a more comprehensive and real-time picture of oral health burdens (Cohen et al., 2024; Daughton, 2020). If a strong correlation between wastewater bacterial levels and oral disease prevalence is established, this approach could serve as an early warning system for emerging trends in oral health conditions, prompting timely public health interventions (Kasprzyk-Hordern et al., 2022; Mao et al., 2020). However, there is no current public health surveillance system for the oral diseases caused by these specific dominant pathogens at the national or local level. Hence, our current understanding on the prevalence of these diseases within the past 5 years has been severely limited. Therefore, the use of WBE is especially timely in filling the gaps in information about these oral pathogens.

Beyond monitoring, the detection of oral bacteria in wastewater raises concerns about their environmental persistence and potential transmission pathways (Chopra et al., 2020; Galarde-López et al., 2024). Although wastewater treatment processes are designed to remove or reduce microbial contaminants, some pathogens can survive or be reintroduced into the environment through treated effluent and biosolids by attaching to particles (Chahal et al., 2016; Marutescu et al., 2023). If *S. mutans* and *P. gingivalis* demonstrate resilience through treatment processes, there is a risk of environmental exposure, which could have implications for public health (Huang et al., 2024; Li et al., 2022; Rams et al., 2023; Sweeney et al., 2004; Teoh et al., 2018). Further studies are needed to assess whether these bacteria can colonize environmental surfaces, influence microbial ecosystems, or contribute to antimicrobial resistance.

In this study, we determined the concentration of *S. mutans* and *P. gingivalis* in wastewater samples over one year. We also determined the concentration of *S. mutans* and *P. gingivalis* in wastewater primary influent, primary effluent, and secondary effluent. Lastly, we observed the prevalence and persistence of *S. mutans* over time in influent wastewater by running a month-long decay experiment. By measuring the prevalence of *S. mutans* and *P. gingivalis* in untreated wastewater and evaluating their reduction through treatment processes, this study aims to bridge a significant knowledge gap in WBE. If successful, integrating oral bacteria surveillance into WBE frameworks could revolutionize how we monitor, predict, and mitigate oral health diseases at a population level. Public health agencies could use wastewater data to design targeted interventions, such as fluoridation programs, oral health education campaigns, or improved access to dental care in communities with high oral bacterial loads (Melbye and Armfield, 2013).

## 2.0 Materials and Methods

### Study site & temporal sampling

100 sewage samples were collected from Blucher Poole Wastewater Treatment Plant, a local wastewater treatment plant (WWTP) in Bloomington, Indiana. Blucher Poole has a flow of more than 4.5 million gallons per day and caters to the northern part of Monroe County. This treatment plant was selected to represent the majority of the Bloomington, Indiana, population and to observe bacterial shedding rates in wastewater. These samples were collected weekly starting from June 2023 to May 2025. A second local WWTP was selected, Dillman Road Wastewater Treatment Plant, so that we were able to compare and contrast between the bacterial quantification of the two plants. Dillman Road is located on the south side of Bloomington, Indiana and has a peak hydraulic capacity at 30 million gallons daily (Bloomington, 2023). For this WWTP, 13 weekly sewage samples from December 2023 to February 2024 were collected and transported to the lab. 24-hour composite influent (prior to primary treatment) wastewater samples were collected in 500-milliliter sterile polypropylene bottles and transported in a cooler and frozen to -20°C within 2 hours of collection. Once received, these samples are immediately prepared for filtration.

### Effluent Sampling

Composite samples of primary and secondary/final effluent wastewater were collected from the local WWTP and placed at -20°C for storage until sample processing. These samples were collected during a 2-week intensive study and in the month of February when concentrations were highest. For the primary influent, a 500 mL 24-hour composite sample was collected, this is the sewage sample that comes directly into the treatment plant and has not received any treatment. We received a 100 mL sample of the 24-hour composite primary effluent sample, which consists of the sewage sample after primary sedimentation has occurred. For the 24-hour composite secondary/final effluent, 500 mL was collected in 500 mL sterile polypropylene bottles. This sample is the sewage sample post aeration, activated sludge, and secondary sedimentation. All samples were transported in the same manner as mentioned previously.

### Decay Experiments

A decay experiment was conducted on Streptococcus mutans in wastewater over the course of a month to observe its persistence. The experiment was conducted in triplicates of 400 milliliters of composite influent wastewater in 500-milliliter flasks. Flasks were then covered with parafilm and left in the dark for a month. 25 mL samples were taken at each of the following time points: 2 hours, 1 day, 4 days, 7 days, 11 days, and 27 days. Before taking each sample, the flasks were swirled to homogenize the wastewater and bacteria inside. Samples were then processed using the methods below and quantified for *S. mutans*.

### Concentration and DNA Extraction

For final effluent samples, 250 mL of the final effluent was taken to be filtered and for all other samples, 50 mL was prepared to be filtered. All samples were acidified to a pH of 3.5. A known concentration of bovine coronavirus solution was also added to each sample as a tracer. For filtration, an electronegative mixed cellulose ester (MCE) 47mm diameter and 0.45 µm pore size filter was used to filter samples based on previous studies (Bivins et al., 2020b; Greaves et al., 2022; Rodriguez et al., 2024). Once filtration through the filter was complete, the filters were then transferred into 2.0 mL PowerBead tubes and either further extracted immediately or stored at -20 °C for extraction later. The DNA from the samples was then extracted in accordance to the provided manufacturer instructions using the Allprep PowerViral DNA/RNA extraction kit (Cat. No. 28000-50 Qiagen, Germantown, MD,USA). This kit resulted in the elution of DNA in 100 µL of RNase-free water provided within the kit. Once eluted these samples were then stored at -20°C in preparation for quantification.

### Bacterial and Viral Quantification

Bacterial quantification of the samples was performed using digital polymerase chain reaction (dPCR) and the QIAcuity™ Four Platform System (Cat. No 911042, Qiagen, Germantown, MD, USA), the QIAcuity Nanoplate 26k (Cat. No 250001, Germantown, MD, USA), and the QIAcuity™ Probe PCR Kit (Cat. No. 250132 Qiagen, Germantown, MD, USA ) as described in previous studies (Rodriguez et al., 2024). The environmental mix for each molecular testing consisted of 1X Qiacuity Probe PCR mix, 08 µM of each forward and reverse primer, 0.4 µM of each probe, and 10 µL of the target DNA for a total volume of 40 µL. The molecular primers and probes used for the detection of *S. mutans* and *P. gingivalis* are described in **Table S1**. While the primer and probe sets were chosen from peer-reviewed, previously validated assays, their specificity was formally assessed through in silico screening using NCBI BLAST. Primer and probe sets were also able to detect a diluted culture of each bacteria. All samples were tested using a duplex, where *S. mutans* was detected in the green channel with the FAM probe and *P. gingivalis* was tested in the yellow channel with the HEX probe. The primers and probes were manufactured by IDT (Coralville, IA, USA). The Zen^TM^ internal quencher (IDTDNA.com) and the 3’ Black Hole ^TM^ quencher were used to double-quench the probes. PCR was run using the following conditions: an initial denaturation step at 95 °C for 2 minutes then 45 cycles of 95 °C for 5 seconds and 60 °C for 30 seconds. The lowest detection limit was estimated for each PCR assay using DNA controls and a dilution series which resulted in a calculated 380 copies/L for both *S. mutans* and *P. gingivalis*. CrAssphage was used as a molecular fecal indicator and quantified with techniques, probes, and primers published previously (Bivins et al., 2020a; Greaves et al., 2020a; Wu et al., 2020) The threshold for each PCR reaction was marked by the top of the negative band of the negative control for the specific probe being tested.

### Data analysis and statistics

The following formula was used to calculate the concentration of oral bacteria in gene copes/L:

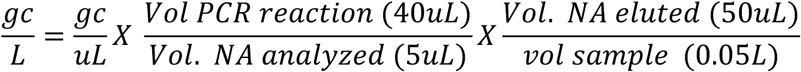

The analyses for our data was conducted using ANOVA and was performed on GaphPad Prism v 10 (Boston, MA, USA) where independent factors were taken into consideration. Graphs and figures were made using GraphPad Prism v 10 (Boston, MA, USA). **For calculating the rate of decay, the decay curve was fit to the following formula:**

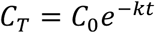

In this equation, C_T_ represents the concentration of the pathogen at time *T*, *C_0_* represents the initial concentration of the pathogen at time 0, and *k* is representative of the decay rate.

## 3.0 Results

### Bacterial and Viral Detection and quantification

**Figure 1** shows the average monthly wastewater flow rates into the main local wastewater treatment plant tested in this study. Values throughout the year show the lowest value being in the summer and the highest values in the winter months. In addition to monitoring flow rates, the fecal indicators crAssphage was measured in all the samples throughout the duration of the experiments and concentrations remained stable over the two year period (**Figure S1**). CrAssphage had an average detected concentration of 8.50 log_10_ GC/L. Statistical tests were done using a one-way ANOVA followed by tukey multiple comparisons to measure changes in monthly concentrations crAssphage, and results showed no statistical differences between crAssphage concentrations over the different months of the experiment.

**Figure 1.**
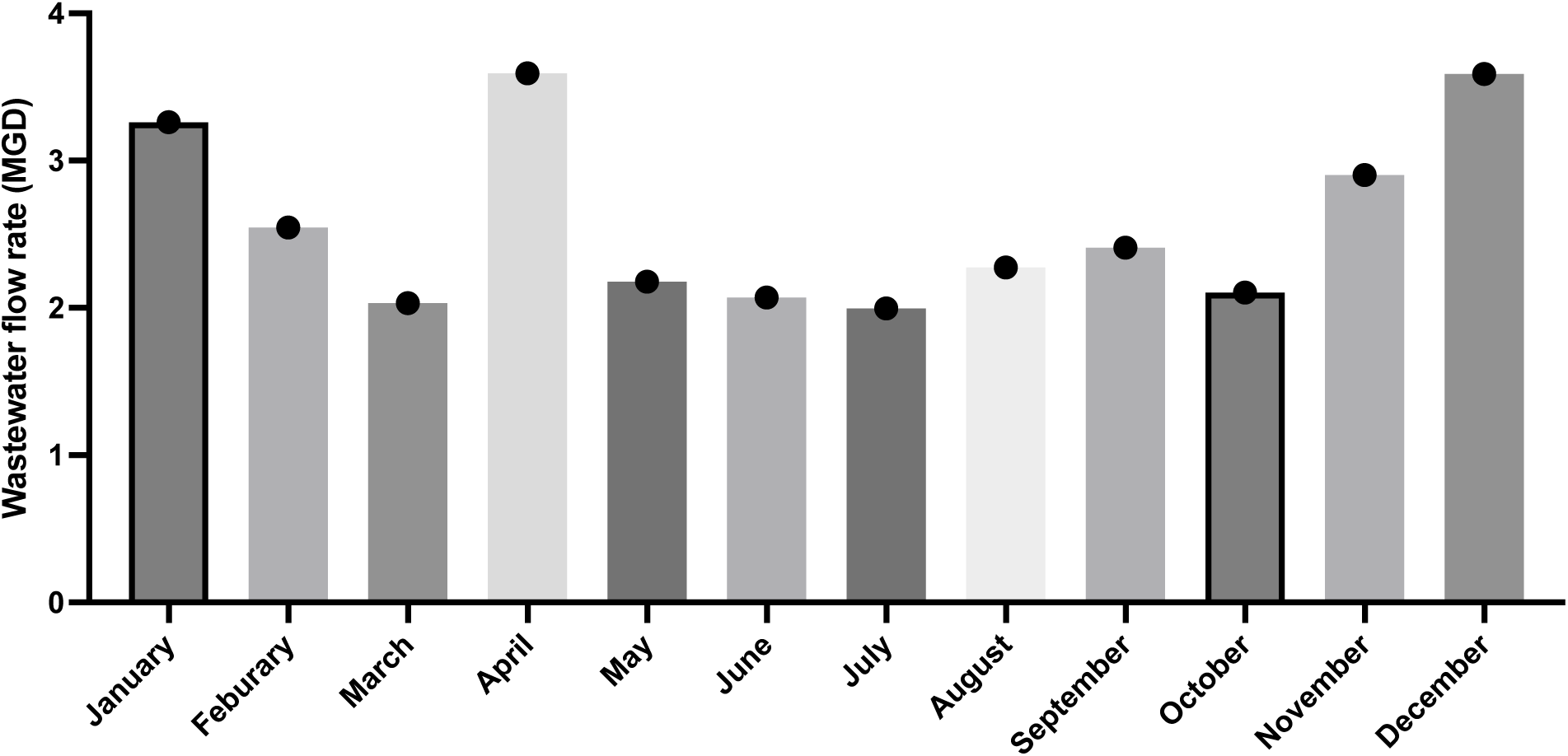
Average wastewater flow rate per month for the local wastewater treatment plant monitored.

**Figure 2** shows the monthly average concentrations over the span of two years for *S. mutans* and *P. gingivalis* in the main local wastewater treatment plant. Bacterial concentrations were normalized by wastewater flow rates to accommodate for population changes. The highest concentration of *S. mutans* was observed in February of year 1 with a calculated monthly average concentration of 5.01 log_10_ GC/L and the lowest detected concentration being in August of year 2 with 3.39 log_10_ GC/L. For *P. gingivalis*, the month with the highest average concentration was November 2023 with a calculated average monthly concentration of 3.81 log_10_ GC/L and the month with the lowest values was September 2024 where none of the samples had detections and the values were assumed to be the detection limit. Over the course of the entire selected timeline, the average detected concentration of *S. mutans* and *P. gingivalis* was 4.57 log_10_ GC/L and 3.03 log_10_ GC/L respectively. **Table 1** breaks down the concentration values for *S. mutans* and *P. gingivalis* into four different time periods within the year, corresponding with Spring (March-May), Summer (June-August), Fall (September-November), and Winter (December-February). Values are also the average of the two years. For *S. mutans* the highest average detected concentration was in the winter with a value of 4.78 log_10_ GC/L and a positive detection rate of 100%. Statistical tests showed winter concentrations were significantly higher than summer and spring concentrations for *S. mutans*. *P. gingivalis* also had the highest average detected concentration in the winter with a value of 3.12 log_10_ GC/L and a positive detection rate of 84% which was also the highest detection rate across all seasons. No significant differences were found between the concentrations across all seasons for *P. gingivalis*.

**Figure 2.**
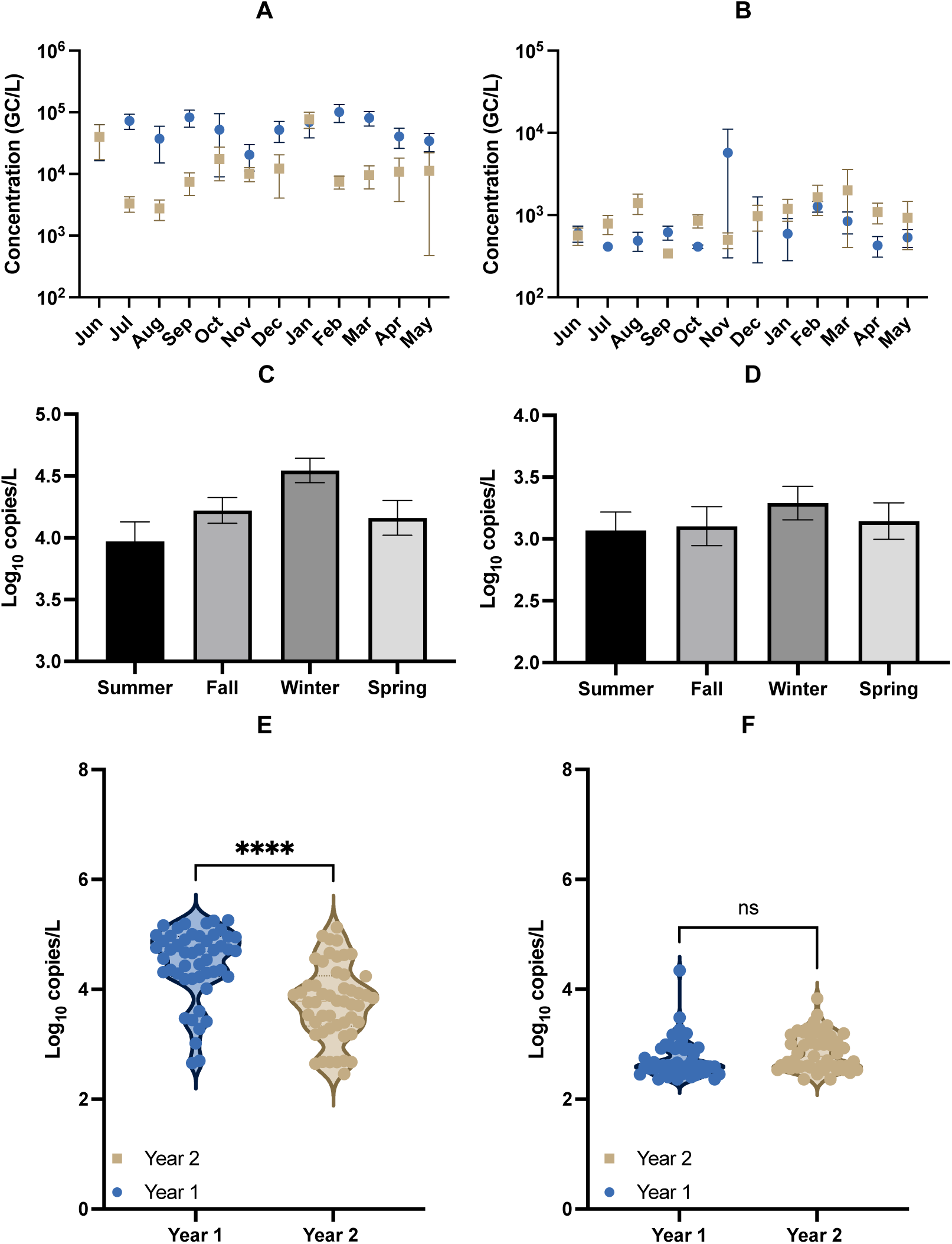
Concentrations of oral bacteria in wastewater. Top two figures show monthly average concentrations of *S.mutans* (A) and *P. gingivalis* (B) in Bloomington wastewater over the two-year duration normalized by wastewater flow. The bars are standard deviation for four samples collected each month (n=4-5). Middle figures show average concentrations for *S.mutans* (C) and *P. gingivalis* (D) in each season over the two years. Bottom figures show violin plots for both oral bacteria in year 1 and year 2 (E for *S.mutans* and F for *P. gingivalis*).

**Table 1:** Detection rates and average concentrations of *S. mutans* and *P. gingivalis* in wastewater samples.

| Target | Seasons | Sample period | No. of Positive Samples (%) | Concentration (Log<sub>10</sub> copies/L) |
| --- | --- | --- | --- | --- |
| S. mutans | Spring 2024 & 2025 | March-May | 19 (76) | 4.37 |
|  | Summer 2023 & 2024 | June-August | 20 (80) | 4.34 |
|  | Fall 2023 & 2024 | Sept-Oct | 25 (100) | 4.35 |
|  | Winter 2024 & 2025 | Jan-Feb & Dec previous year | 25 (100) | 4.78 |
|  | Total |  | 89 (89) | 4.57 |
| P. gingivalis | Spring 2024 & 2025 | March-May | 12 (48) | 2.93 |
|  | Summer 2023 & 2024 | June-August | 12 (48) | 2.75 |
|  | Fall 2023 & 2024 | Sept-Oct | 13 (52) | 3.04 |
|  | Winter 2024 & 2025 | Jan-Feb & Dec previous year | 21 (84) | 3.12 |
|  | Total |  | 58 (58) | 3.03 |

A comparison was also done between year 1 (June 2023 to May 2024) and year 2 (June 2024 to May 2025) through using Pearson’s correlation r. Statistical analysis showed no correlation between the different years for both bacteria (**Figure S3**). Correlation analysis for crAssphage for the different years showed a strong positive correlation (r=0.77). A comparison was made between two different WWTP, Blucher Poole (BP) and Dillman Road (DR), for the two bacteria in the winter of year 2, which can be observed in **Figure S2**. For both WWTPs, *S. mutans* was observed to have a larger spread in concentrations than *P. gingivalis* which can be observed in the top two graphs within **Figure S2**. More samples of *P. gingvalis* were disregarded due to not being detected, in comparison to *S. mutans*. For BP, the average concentration of *S. mutans* was 4.93 log_10_ GC/L and for *P. gingivalis* it was 3.14 log_10_ GC/L. The average concentration of *P. gingivalis* for DR was higher at 3.53 log_10_ GC/L whereas *S. mutans* was relatively similar to BP with an average concentration of 4.86 log_10_ GC/L. *P. gingivalis* was statistically higher in the DR WWTP compared to the BP WWTP whereas there were no statistical differences between the concentrations of *S. mutans* in the DR and BP WWTPs.

### Effluent sampling

In **Figure 3** the concentrations of *S. mutans* and *P. gingivalis* in primary influent, primary effluent, and secondary effluent are shown. For both species of oral bacteria, they were most prevalent in the primary influent samples with *S. mutans* having an average concentration of 3.87 log_10_ GC/L and for *P. gingivalis*, a concentration of 3.21 log_10_ GC/L. The difference in concentration between the primary influent and the primary effluent samples was larger for *P. gingivalis* than it was for *S. mutans* with calculated differences of 1.08 log_10_ GC/L and 0.106 log_10_ GC/L, respectively. The final effluent samples still resulted in some detected concentrations, with *S. mutans* having an average of 3.08 log_10_ GC/L and *P. gingivalis* having an average of 1.99 log_10_ GC/L. The difference between primary influent and secondary effluent for both *S. mutans* and *P. gingivalis* was significant (p<0.0001). For *S. mutans*, the average concentration for primary effluent and secondary effluent was determined to be significant as well (p<0.0001). The difference between the concentration of *P. gingivalis* in primary influent and primary effluent samples was significant (p=0.009). There was determined to be no significance in the concentration of *S. mutans* in the primary influent and the primary effluent samples (p=0.816) as well as there being no significant difference in the concentration of *P. gingivalis* between the primary effluent and secondary effluent samples (p=0.064).

**Figure 3.**
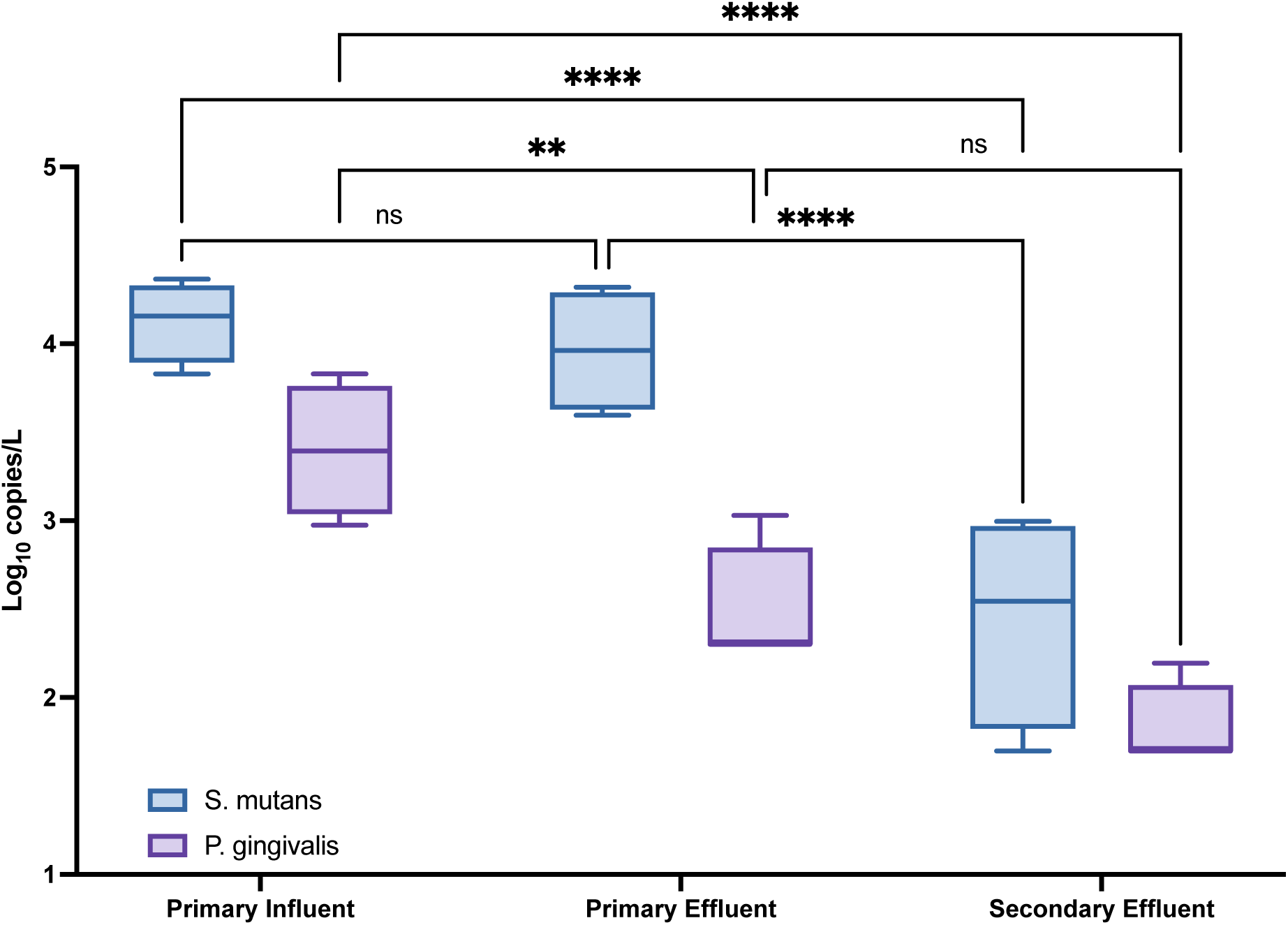
Occurrence of *S. mutans* and *P. gingivalis* in wastewater treatment plant primary influent, primary effluent, and secondary effluent in 4 replicate samples.

**Figure 4.**
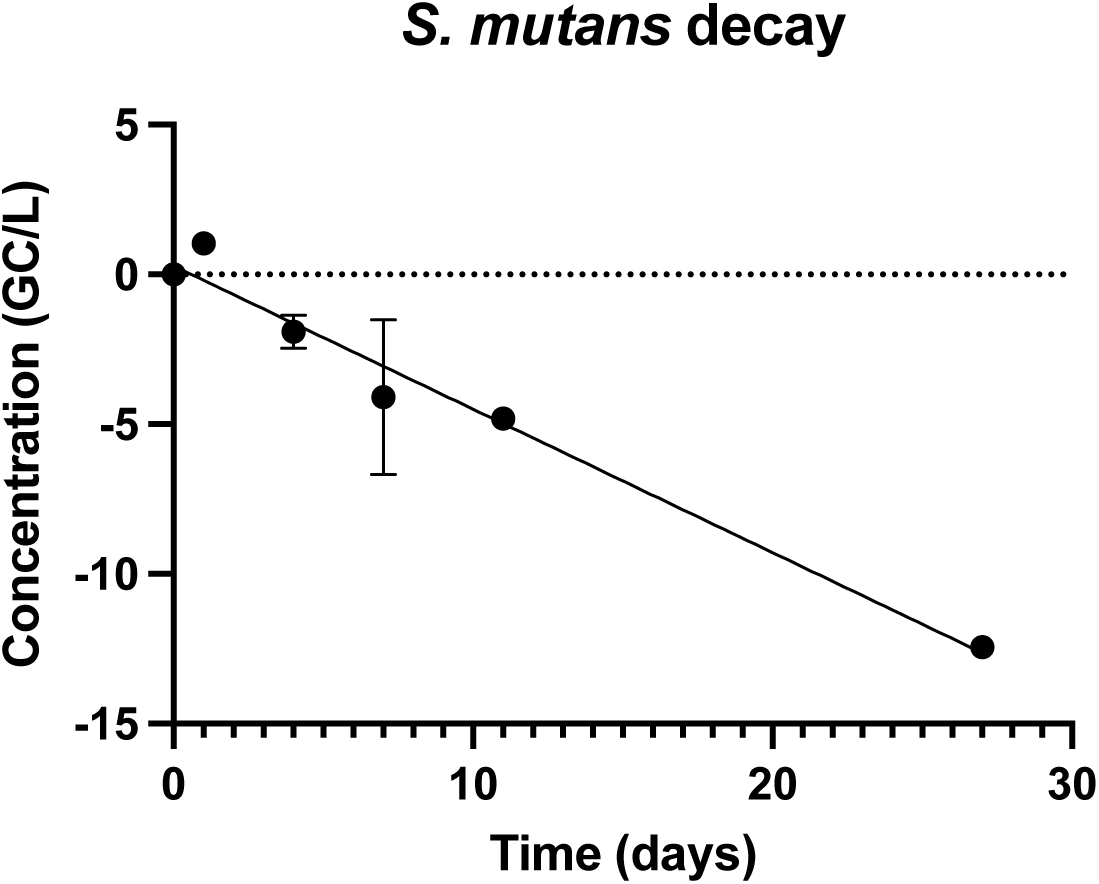
Decay of *S. mutans* in wastewater primary influent at room temperature in the absence of sunlight.

### Decay Rate

The decay rate of *S. mutans* at room temperature in the absence of sunlight was observed over the span of one month, with a visual representation of the data being shown in **Figure 3**. There was positive detection for *S. mutans* all the way up to 27 days with an average value of 4.05 log_10_ GC/L. Across all three replicates, the average *k_obs_* for *S. mutans* was -0.4786 d^-1^. For *P. gingivalis*, the decay rate was unable to be observed due to the low initial concentration of the bacteria in wastewater.

## 4.0 Discussion

To the best of the authors’ knowledge, this study is the first study to detect and quantify *S. mutans* and *P. gingivalis* in wastewater samples. These two pathogens cause two of the most common dental diseases that infect a majority of populations worldwide. Even though these pathogens are highly prevalent in US populations, routine surveillance on the prevalence of these different pathogens or the oral disease they cause is not done, especially since the outbreak of the COVID-19 pandemic (Dani et al., 2016; Yan et al., 2021). Through our study, we were able to show that both pathogens were detected in more than 60% of the 65 samples tested in wastewater over the 4 different seasons in a year. The high detection of these oral pathogens in wastewater samples from a mid-size city (75,000 residents) shows that wastewater-based epidemiology (WBE) is highly feasible for these dental pathogens and oral health. WBE may also serve as a low-cost method to monitor the spread and prevalence of these dental diseases when traditional monitoring methods are not done.

*S. mutans* had consistently higher concentrations than *P. gingivalis* in wastewater throughout the sampling period. In some samples, concentrations of *S. mutans* reached to more than 10 times higher than *P. gingivalis*. With numerous studies estimating higher percentages of tooth decay (90%) than gum disease (50%) in adults (Research, 2022; United States. Public Health Service. Office of the Surgeon General and National Institute of Dental and Craniofacial Research (U.S.), 2021), our differential observations of *S. mutans* (which associates with tooth decay) and *P. gingivalis* (which associates with gum disease) is consistent with past clinical prevalences of these diseases in US populations (Griffen et al., 1998; Rafiei et al., 2018; Thorild et al., 2002; Zubaidah et al., 2022). Higher prevalences of a disease should result in higher concentrations in wastewater (Rabe et al., 2023; Weidhaas et al., 2021). However, concentrations of *S. mutans* could also be higher in wastewater due to the shedding dynamics from teeth as opposed to the shedding dynamics of *P. gingivalis* from gums (Bugueno et al., 2020; Liao et al., 2014; Liu et al., 2017). While studies have not specifically tested shedding of each pathogen, previous studies have observed statistically higher concentrations of *S. mutans* than *P. gingivalis* in dental samples from healthy patients, alluding to higher shedding of *S. mutans* than *P. gingivalis* (Almeida et al., 2020; Dani et al., 2016; Davis et al., 2012). Differential shedding could be due to the stark difference in oral hygiene when it comes to the gums and teeth where cleaning of the teeth is done more frequently than cleaning of the gums (Albertsson and Dijken, 2010; Deinzer et al., 2019; Goryawala et al., 2016; Zimmermann et al., 2015). The variations in shedding might also be simply attributed to the ease with which they attach or detach from different surfaces, such as enamel versus gums (Gibbons, 1984; Mei et al., 2009; Vanhoute, 1983).

In addition to being shed in saliva, oral pathogens may also be swallowed, survive passage through the gastrointestinal tract, and be excreted in feces. Several studies have detected the DNA of oral microbes, including S. mutans, in human feces, particularly through 16S rRNA gene sequencing (Abdelbary et al., 2022; Lin et al., 2024). This fecal shedding pathway may significantly influence the concentration of these bacteria in wastewater. For example, *S. mutans* may be more resilient than *P. gingivalis* during gastrointestinal transit, potentially resulting in higher levels of S. mutans in fecal matter. While feces are generally considered the primary route of microbial contribution to wastewater, no dedicated studies have confirmed the specific shedding patterns of *S. mutans* and *P. gingivalis* (Newton et al., 2015). As a result, there remains uncertainty about how their wastewater concentrations correlate with actual infection prevalence in the population. Future studies should examine the shedding dynamics of each oral pathogen so that improved connections between dental health and wastewater concentrations can be developed.

### Seasonal Patterns

WBE has been shown to be a very useful tool for understanding disease trends, specifically over the different seasons of a year (Brinkman et al., 2017; Kuhn et al., 2023). When trends for different seasons are determined, health agencies can then pick specific seasons throughout the year to focus mitigation efforts on (dos Santos et al., 2024; Mao et al., 2020). Our results showed seasonal trends for *S. mutans* and *P. gingivalis,* with both oral bacteria having their highest average concentrations in the winters over the two-year period. However, results for *P. gingivalis* were not significant, potentially due to the low detection rate of this pathogen. Nevertheless, detections in winter for *P. gingivalis* were also much higher than the detections in other months, suggesting potential seasonality similar to *S. mutans*.

*S. mutans* showed significantly higher concentrations in the winter than in the spring and summer. The highest temperature months of July and August exhibited notably lower concentrations compared to the other months of the year, especially in year 2. These results highlight that *S. mutans* and dental caries could be more prevalent in months with colder weather. Prior studies have observed diminished oral health in colder months due to drier mouths and immune system changes in the winter (Fares, 2013; Fisman, 2007). Drier mouths due to increased indoor heater use can reduce the volume of saliva (known to prevent bacterial growth), which in turn can cause the extended growth of pathogens like *S. mutans* (King et al., 2000; Ligtenberg et al., 2020; Weng et al., 2022). Colder months can also lead to diet changes, lowering nutrient levels, or less hydration which could potentially contribute to tooth decay and poorer oral health in colder months compared to hotter months (Fares, 2013; Moynihan and Petersen, 2004; Sheiham, 2001). Although we observed higher average concentrations of oral pathogens during the winter months across both years, several outlier months in each season deviated from this trend, suggesting that additional factors may be influencing oral health dynamics. These findings are also region-specific; other areas may exhibit different seasonal patterns for these two oral pathogens. Future studies should integrate oral health indicators with behavioral and environmental factors to uncover the underlying drivers of seasonal variation.

Though average concentrations in the different seasons across years show potential seasonal trends, trends of bacteria in each year were not well correlated (**Figure S3**). Specifically for S. mutans, our data observed significantly lower concentrations in year 2 than in year 1 suggesting reduction in oral pathogen burden in the community. P. gingivalis has statistically similar concentrations across both years but concentrations over time were also not well correlated. A potential reason for reduction in oral health burden is that COVID-19-related dental clinic closures and delayed care likely worsened oral health, but as services resumed, individual dental health may have improved, potentially lowering oral health-related markers in wastewater over time (Brian and Weintraub, 2020; Dill et al., 2023). Prior clinical studies have also showed lowering oral health burden over the past decade which follow the trend in our study (Dai et al., 2025; Lagerweij and van Loveren, 2015; Qin et al., 2022). Additionally, year to year patterns can significantly change depending on healthcare access, economic conditions, and public health policies that affect community oral health in a year (Cooray et al., 2025; Dunleavy et al., 2024; Lee et al., 2021).

### Variation in WWTP

Our study results showed that *S. mutans* concentrations in wastewater from both WWTP’s were not statistically different within the wintertime even though both WWTPs serve different population numbers and types. These results suggest that *S. mutans* can be prevalent in different WWTP and populations across geographical areas. The higher observed concentration of *S. mutans* compared to *P. gingivalis* in the DP WWTP is consistent with what was observed for BR WWTP. Our results do show statistically higher concentrations of *P. gingivalis* in the DR WWTP compared to the BP WWTP (p=0.0273). BP WWTP primarily serves the university campus community, which is mostly comprised of young students between the ages of 18 and 24. DR WWTP serves the rest of the local population which contains children and vulnerable groups that may not be present in the other WWTP. As oral health significantly impacts those that are most vulnerable, such as children (below 18) and elderly (above 65), it is expected that there would be higher levels of reduced oral health in the DR WWTP which contains these vulnerable groups (Bloomington, 2023; Weng et al., 2022; Xiao et al., 2020). However, to specifically determine whether these factors affect oral health, further analysis is needed on samples across the sewershed and in other geographic locations.

### Fate in the wastewater treatment process

Throughout the wastewater treatment process, both *S. mutans* and *P. gingivalis* were detectable but at significantly lower concentrations. These data suggest that the treatment process is reducing the oral bacterial population, but not completely. The primary treatment step was able to significantly reduce *P. gingivalis* but the secondary treatment step (both activated sludge and secondary sedimentation process) was needed to reduce *S. mutans*. These differences are potentially due to how likely each microbe is to stick to fecal particles in wastewater. Multiple prior studies have shown a reduction of various pathogens in wastewater through the primary and secondary treatment processes (Delanka-Pedige et al., 2020; Frigon et al., 2013; Fu et al., 2010; Skouteris et al., 2020). The same studies also show that certain pathogens are able to survive through the process and enter the environment at lower levels, similar to what we are observing for *P. gingivalis* and *S. mutans* (Delanka-Pedige et al., 2020; Frigon et al., 2013; Fu et al., 2010; Skouteris et al., 2020) (Cuevas-Ferrando et al., 2022). Beyond assessing the fate of oral bacteria in the wastewater treatment plant, our study assessed the decay of oral bacteria in wastewater. With *P. gingivalis* having very low concentrations, assessing its decay in water would be difficult. Furthermore, as *P. gingivalis* is an anaerobic bacteria, its persistence in wastewater would be lower than *S. mutans* due to the presence of oxygen in wastewater (Diaz and Rogers, 2004; Lemos et al., 2019; Rafiei et al., 2018; Skouteris et al., 2020). Our calculated decay rates (0.48 d^-1^) for *S. mutans* in wastewater are comparable with other types of common fecal bacteria found in wastewater and the environment, such as E. coli (-0.14 to -1.13 day^-1^) (Chan et al., 2015; Nakhle et al., 2021; Tiwari et al., 2023) and enterococci (-0.19 to -1.2 day^-1^)(Greaves et al., 2020b; Sagarduy et al., 2019; Tiwari et al., 2019; Tiwari et al., 2023). This environmental and wastewater treatment plant survival, therefore, highlight the potential novel transmission pathway of these oral pathogens through the environment in contaminated waters.

### Limitations

A key limitation of our study is the inability to compare our wastewater data with clinical diseases that these pathogens cause (periodontal diseases and dental caries). The main reason why we cannot access this data is because there is no clinical surveillance of these pathogens in our region which highlights how important wastewater surveillance can be to the monitoring of oral health. Future studies should try to incorporate clinical studies or survey data with wastewater surveillance to better connect wastewater data with the oral health of the community. Another major limitation includes the use of only molecular methods to detect the various oral pathogens in this study. While molecular methods offer fast and precise measurements for a diverse range of targets, they do not assess viability of specific organism. Subsequent studies should incorporate culturable or other viability assessment methods to measure the viable concentration of oral pathogens, especially when investigating persistence.

## 5.0 Conclusion

In conclusion, this study demonstrated the feasibility of using WBE for oral pathogen surveillance in a community. Additionally, our study showed a potential environmental transmission pathway for *S. mutans* through water. Overall, this research has the potential to provide critical insights into oral health trends, improve disease surveillance, and enhance public health strategies. By leveraging wastewater analysis, we can move toward a more proactive and non-invasive approach to oral disease monitoring, ultimately contributing to better health outcomes and reducing the global burden of dental and periodontal diseases.

## Data Availability

All relevant data are within the manuscript and its Supporting Information files.

